# Comparison of Stress-rest and Stress-LGE analysis strategy in patients undergoing stress perfusion cardiovascular magnetic resonance

**DOI:** 10.1101/2023.08.08.23293861

**Authors:** Peter P. Swoboda, Gareth Matthews, Pankaj Garg, Sven Plein, John P. Greenwood

**Author notes:** **Corresponding author: John P. Greenwood** MB ChB, PhD, Multidisciplinary Cardiovascular Research Centre and Department of Biomedical Imaging Science, Leeds Institute of Cardiovascular and Metabolic Medicine, University of Leeds, Leeds LS2 9JT, United Kingdom.

## Abstract

**BACKGROUND:** Stress perfusion cardiovascular magnetic resonance (CMR) is increasingly used without rest perfusion for the quantification of ischemia burden. However, the optimal method of analysis is uncertain.

**METHODS:** We identified 666 patients from Clinical Evaluation of MAgnetic Resonance imaging in Coronary heart disease (CE-MARC) with complete stress perfusion, rest perfusion, late gadolinium enhancement (LGE) and quantitative coronary angiography (QCA) data. For each segment of the 16-segment model, perfusion was visually graded during stress and rest imaging, with infarct transmurality assessed from LGE imaging. In the “Stress-LGE” analysis a segment was defined as ischemic if it had a subendocardial perfusion defect with no infarction. Rest perfusion was not used in this analysis. We compared the diagnostic accuracy of “Stress-LGE” analysis against QCA and the “Stress-rest” method validated in the original CE-MARC analysis. The diagnostic accuracy of the “Stress-LGE” method was evaluated with different thresholds of infarct transmurality, used to define whether an infarcted segment had peri-infarct ischemia.

**RESULTS:** The optimal “Stress-LGE” analysis classified all segments with a stress perfusion defect as ischemic unless they had >75% infarct transmurality (AUC 0.843, sensitivity 75.6%, specificity 93.1%, P<0.001). This analysis method has superior diagnostic accuracy to the “Stress-rest” method (AUC 0.834, sensitivity 73.6%, specificity 93.1%, P<0.001, P-value for difference=0.02). Patients were followed up for median 6.5 years for major adverse cardiovascular events (MACE), with the presence of inducible ischemia by either the “Stress-LGE (>75%)” or “Stress-rest” analysis being similar and strongly predictive (Hazard Ratio 2.65, P<0.0001, for both).

**CONCLUSIONS:** The optimum definition of inducible ischemia was the presence of a stress-induced perfusion defect without transmural infarction. This definition improved the diagnostic accuracy compared to the “Stress-rest” analysis validated in CE-MARC without the need for rest perfusion imaging. The absence of ischemia by either analysis strategy conferred a favorable long-term prognosis.

**CLINICAL PERSPECTIVE:** *What is new?:* Functional ischemia testing, specifically with stress perfusion cardiovascular magnetic resonance (CMR), is an established step in the evaluation of patients with chest pain. This study demonstrates that the rest perfusion imaging can safely be removed from the acquisition and analysis, without compromising imaging diagnostic and prognostic accuracy. For the highest diagnostic accuracy, all segments with stress-induced subendocardial hypoperfusion without transmural infarction should be considered ischemic.

*What are the clinical implications?:* Removal of rest imaging from the stress perfusion CMR examination reduces study duration which could potentially reduce costs, increase throughput, and build capacity to increase access to CMR.

## INTRODUCTION

Stress perfusion cardiovascular magnetic resonance (CMR) has a Class 1 recommendation in the 2021 AHA/ACC/ASE/CHEST/SAEM/SCCT/SCMR guidelines for the evaluation and diagnosis of chest pain, particularly in those with intermediate-high risk or known obstructive coronary artery disease^1^. Conventionally perfusion imaging is performed first during stress with a short acting vasodilator such as adenosine, then again at rest, with segmental hypoperfusion at stress that normalizes at rest being diagnostic for ischemia.

Society for Cardiovascular Magnetic Resonance (SCMR) Standardized Protocols recommend that a minimum of 10 minutes should be left between stress and rest imaging to ensure the hemodynamic effects of adenosine have resolved^2^. A further 5-minute gap between rest perfusion and late gadolinium enhancement (LGE) imaging is advised. Removing rest perfusion imaging from the protocol can therefore save at least 5-7 minutes from the scan, and the standardized protocols advise that this can be done depending on institutional policy and experience.

The stress-rest ischemia definition has been validated in several studies including Clinical Evaluation of MAgnetic Resonance imaging in Coronary heart disease (CE-MARC), a prospective study of 752 patients who underwent stress perfusion CMR, myocardial perfusion scintigraphy using single-photon emission computed tomography (MPS-SPECT) and invasive coronary angiography^3^. In this study, ischemia, defined as a myocardial perfusion defect seen during hyperemia but not at rest, was found to have sensitivity of 77% and specificity of 92% compared to quantitative invasive angiography for the detection of significant coronary artery disease (CAD). A major strength of CE-MARC was that all patients underwent both CMR and invasive angiography, and that the CMR data was not used in clinical decision making.

There has been a recent trend to perform a stress-only protocol in which inducible ischemia is deemed to be present if there is a perfusion defect on stress perfusion imaging but no scar detected on LGE imaging^2,4,5^. Although removal of rest perfusion from the CMR exam has the potential to shorten the overall study time, its diagnostic accuracy against invasive coronary angiography is not known.

We hypothesized that when interpreting stress perfusion CMR, “Stress-LGE” analysis has equivalent diagnostic accuracy to the “Stress-rest” analysis that was validated in the original CE-MARC study. We also aimed to identify the optimum threshold of infarct transmurality at which segments with both infarction and stress-induced subendocardial hypoperfusion should be considered ischemic.

## METHODS

The study design and primary analyses have been published previously^3,6^. In brief, patients with suspected stable angina were prospectively enrolled if they had at least 1 major cardiovascular risk factor and a cardiologist considered them to require further investigation. By protocol, all patients were scheduled to have CMR and MPS-SPECT in a randomized order, followed by invasive fluoroscopic coronary angiography within 4 weeks, regardless of the treating physician’s chosen clinical pathway or prior imaging test results. After invasive angiography, the MPS-SPECT results could be made available on request to enable decision making about revascularization (blinding the treating clinician to this result was deemed unethical); however, CMR results were kept blinded. The study was conducted in accordance with the Declaration of Helsinki (2000) and approved by the United Kingdom National Research Ethics Service (05/Q1205/126); all patients provided informed written consent. Extended 5-year follow-up was conducted with approval from the National Research Ethics Service (14/YH/0137) and under Section 251 of the National Health Service Act 2006 (14/CAG/1018). Original data and analysis utilized in the study are available from the corresponding author at reasonable request.

### CMR acquisition and analysis

CMR was performed on a 1.5T Philips Intera system (Philips, Best, The Netherlands) using a protocol that included stress perfusion (adenosine, 140μg/kg/min for ≥4 minutes), cine imaging, rest perfusion and LGE. CMR coronary imaging was previously shown to have inferior sensitivity and specificity compared to stress perfusion testing, and was therefore not used in this sub-analysis^3,7–9^. CMR analysis techniques from the original trial, which have previously been described in detail, were used in this analysis^6^. Scans were reported by paired readers with >10 years’ experience who were blinded to other tests and recorded by consensus.

Each segment within the 16-segment model^6^ during perfusion imaging was visually graded at stress and then rest (0 = normal, 1 = equivocal, 2 = subendocardial defect, 3 = transmural defect, 4 = transmural defect and wall thinned). LGE imaging was scored using the same 16- segment model, the presence of any infarction and its pattern described (subendocardial or transmural), and then categorized according to transmurality (0 = normal, 1 = <25%, 2 = 25- 50%, 3 = 50-75%, 4 = 75-100%). Manual co-registration was used to localize infarction and ischemia to each of the 16-segments.

In the “Stress-rest” analysis, as used in the original CE-MARC study, ischemia was defined as an increase in score of ≥2 in at least one segment between stress and rest imaging. LGE imaging was not used in the diagnosis of ischemia in this analysis. For the “Stress-LGE” analysis, a segment was defined as ischemic if it had a stress perfusion score of ≥2 and no infarction (score 0) on corresponding LGE images (Figure 1). Rest perfusion imaging was not used in the diagnosis of ischemia in this analysis. Diagnostic accuracy was assessed by altering the definition of inducible ischemia according to the infarct transmurality (i.e. “Stress-LGE (>25%)”= stress perfusion score ≥2, LGE score ≥1, “Stress-LGE (>50%)”= stress perfusion score ≥2, LGE score ≥2 and “Stress-LGE (>75%)”= stress perfusion score ≥2, LGE score ≥3). Ischemia burden was defined as the number of segments by each method that met the definition for inducible ischemia.

**Figure 1.**
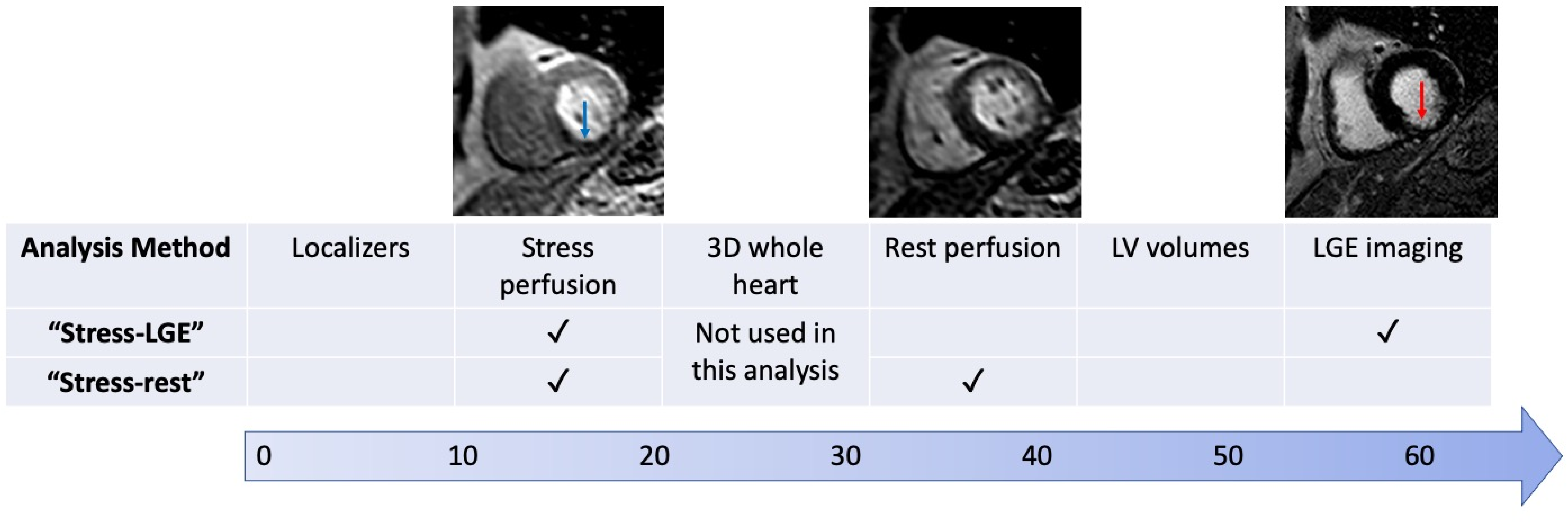
Workflow for “Stress-LGE” and “Stress-Rest” analysis. Arrow denotes number of minutes duration of the scan.

### Invasive coronary angiography

Invasive fluoroscopic coronary angiography was analyzed by two experienced cardiologists blinded to the CMR and MPS-SPECT results. Based on the original trial, significant coronary artery disease was defined as ≥70% stenosis of a first order coronary artery measuring ≥2mm in diameter or left main stem stenosis ≥50% by quantitative coronary angiography (QCA) (QCAPlus, Sanders Data Systems, Palo Alto, California, USA).

### Follow-up

Annual follow-up for 5 years was planned for all recruited patients. A detailed medical history since randomization was obtained from all hospital and general practitioners’ records, then cross-referenced to information obtained by direct telephone contact with each patient. Mortality and cause of death were obtained from the Office for National Statistics via the Health and Social Care Information Centre. Major Adverse Cardiac Events (MACE) was defined as the composite end point of cardiovascular death, myocardial infarction (MI) or acute coronary syndrome, unscheduled coronary revascularization or hospital admission for a cardiovascular cause (stroke/transient ischemic attack, heart failure and arrhythmia), in keeping with previous studies^10,11^. Unscheduled coronary revascularization was defined as any revascularization that occurred owing to clinical deterioration and excluded procedures that were planned based on the index coronary angiography results. All clinical events were adjudicated by a clinical events committee that was blinded to any of the CMR results.

### Statistical analysis

Continuous data are presented as mean ± standard deviation. Diagnostic accuracy of both methods against QCA was assessed by Receiver Operator Curve (ROC) analysis using the method described by Delong^12^. Hazard ratios for MACE were calculated by Cox proportional hazards regression.

## RESULTS

We identified 666 patients from CE-MARC with complete CMR stress perfusion, rest perfusion, LGE and QCA data. By QCA analysis, 262 (39.3%) cases were defined as having significant coronary stenosis.

### Diagnostic accuracy versus angiography

By “Stress-LGE (>0%)” analysis, where all segments with LGE were considered non-ischemic, the area under the curve (AUC) was 0.825 with sensitivity 71.4% and specificity 93.4%. The diagnostic accuracy of “Stress-LGE” analysis could be improved with incremental LGE thresholds: “Stress-LGE (>25%)” AUC 0.835, “Stress-LGE (>50%)” 0.839 and “Stress-LGE (>75%)” AUC 0.843. The diagnostic accuracy of all these definitions was significantly better than “Stress-LGE (>0%)” (Table 1). By “Stress-rest", as used in the original study, AUC was 0.834 with sensitivity 73.6% and specificity 93.1%. The only “Stress-LGE" analysis to have significantly better diagnostic accuracy compared to “Stress-rest” was “Stress-LGE (>75%)” with a difference in AUC of 0.009 (P=0.02). There were no subgroups of patients, including male sex, co-morbidities, prior MI or left ventricular systolic dysfunction (LVSD), where there was any significant difference in the diagnostic accuracy of “Stress-rest” and “Stress-LGE (>75%)” (Table 2).

**Table 1.**
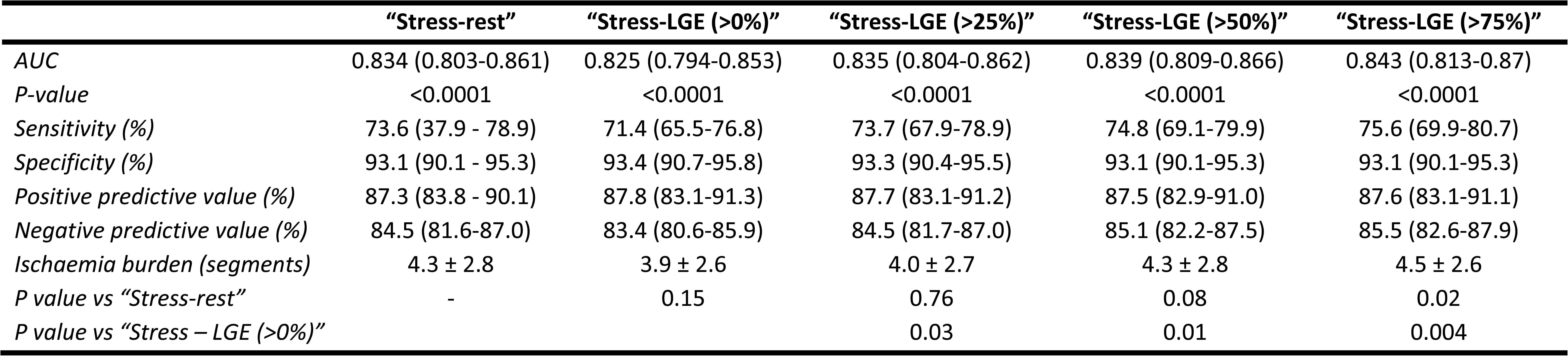
Comparison of stress-LGE analysis methods according to the threshold at which a segment with infarction on LGE can be described as ischaemic. Any segment in the “Stress-LGE (>0%)” analysis with LGE was classified as infarcted and could therefore not be classified as ischemic. In the “Stress-LGE (>75%)” analysis only segments with >75% infarct transmurality were classified as infarcted and segments with 0-75% infarction could be classified as ischaemic if there was inducible ischemia present on stress perfusion imaging. Comparisons in AUC between “Stress-LGE (>25%)”, “Stress-LGE (>50%)” and “Stress-LGE (>75%)” were non-significant.

**Table 2.** Diagnostic accuracy of “Stress-rest” and “Stress-LGE” strategies in subgroups of patients from CE-MARC. SE indicates the Standard Error.

|  | “Stress-rest strategy” |  |  | “Stress-LGE (>75%) strategy” |  | P-value |
| --- | --- | --- | --- | --- | --- | --- |
|  | N | AUC<br>(95% CI) | SE | AUC<br>(95% CI) | SE |  |
| *All patients* | 666 | 0.83<br>(0.80-0.86) | 0.02 | 0.84<br>(0.81-0.88) | 0.02 | 0.15 |
| *Male* | 410 | 0.83<br>(0.79-0.86) | 0.02 | 0.84<br>(0.80-0.87) | 0.02 | 0.25 |
| *Hypertension* | 339 | 0.82<br>(0.78-0.86) | 0.02 | 0.83<br>(0.78-0.87) | 0.02 | 0.52 |
| *Diabetes* | 84 | 0.75<br>(0.65-0.84) | 0.05 | 0.77<br>(0.69-0.85) | 0.05 | 0.66 |
| *Current smoker* | 120 | 0.89<br>(0.81-0.94) | 0.03 | 0.89<br>(0.82-0.94) | 0.03 | 0.75 |
| *Obesity (BMI>30)* | 263 | 0.84<br>(0.79-0.88) | 0.02 | 0.84<br>(0.79-0.89) | 0.02 | 0.66 |
| *Prior ACS* | 55 | 0.8<br>(0.67-0.89) | 0.06 | 0.78<br>(0.64-0.88) | 0.06 | 0.66 |
| *LVEF <50%* | 93 | 0.91<br>(0.83-0.96) | 0.03 | 0.91<br>(0.83-0.96) | 0.03 | 0.95 |
| *LV Hypertrophy*<br>*(LVMI>60g/m<sup>2</sup>)* | 73 | 0.88<br>(0.78-0.94) | 0.04 | 0.89<br>(0.79-0.95) | 0.04 | 0.64 |
| *Any MI on LGE* | 124 | 0.83<br>(0.75-0.89) | 0.05 | 0.86<br>(0.78-0.91) | 0.04 | 0.47 |

### Ischemia Burden

MI was present in 124 (18.6%) patients, where it affected 2.9±2.1 segments. The ischemia burden by “Stress-rest” was 4.3±2.8 segments and increased according to transmurality used in the definition of “Stress-LGE”: for “Stress-LGE (>0%)” 3.9 ± 2.6 segments, “Stress-LGE (>25%)” 4.0 ± 2.7 segments, “Stress-LGE (>50%)” 4.3 ± 2.8 and “Stress-LGE (>75%)” 4.5 ± 2.6 segments (Table 1).

### Patient outcomes

Patients were followed up for a median of 6.8 years, during this time 109 (16.4%) patients suffered at least 1 MACE event. The Hazard Ratio for the presence of inducible ischemia by “Stress-rest”, “Stress-LGE (>0%)” and “Stress-LGE (>75%)” were 2.65, 2.48, and 2.65 respectively (Table 3 and Figure 2) for MACE events (all significant at P<0.001). The presence of inducible ischemia was still associated with MACE by all definitions after correcting for the presence of LGE and the LVEF (Table 4).

**Figure 2.**
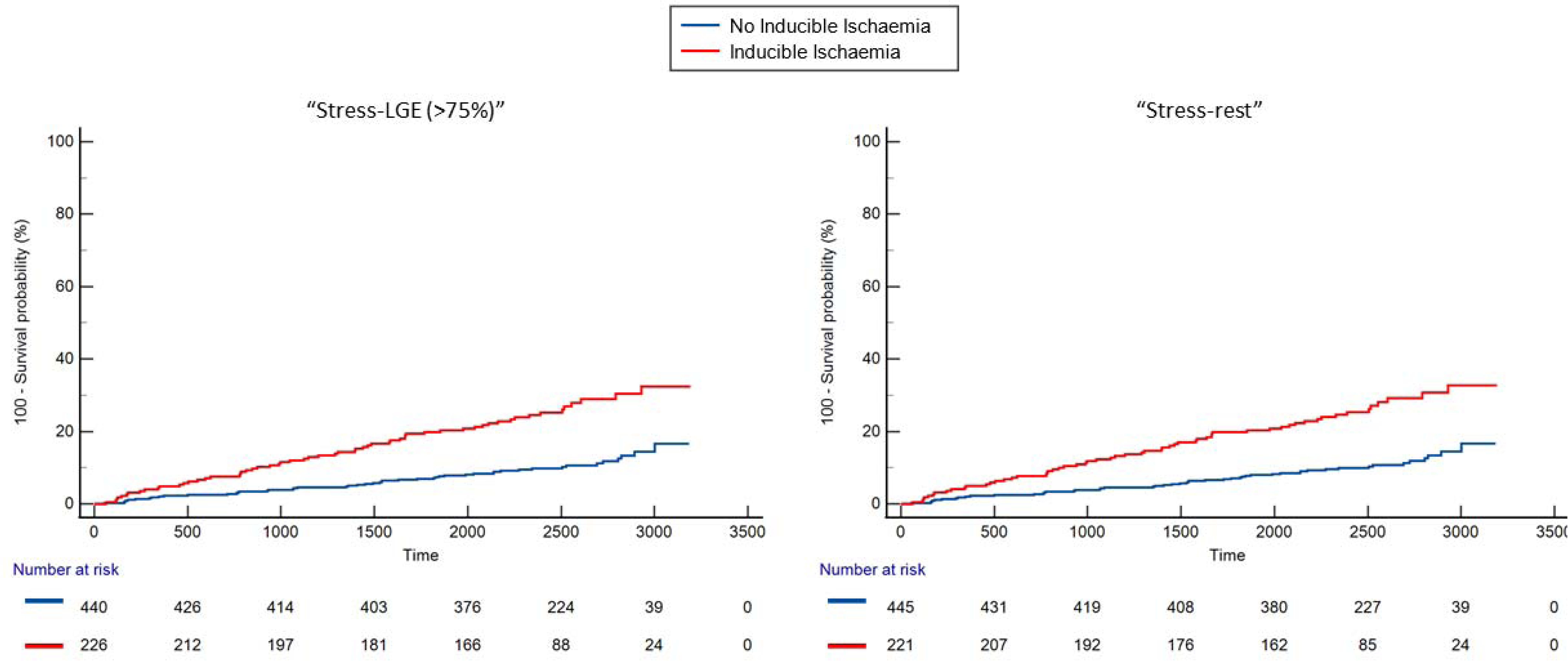
Cumulative MACE according to whether ischaemia is present by “Stress-LGE (>75%)” or “Stress-rest” definitions. P-value by log-rank <0.001 for both.

**Table 3.** Univariable Cox regression of “Stress-rest” and “Stress-LGE” strategies compared to other CMR findings for the prediction of MACE. Beta is the coefficient of the model, SE the Standard Error, and HR the Hazard Ratio.

| Covariate | Beta | SE | HR (95% CI) | P-value |
| --- | --- | --- | --- | --- |
| Any ischemia by “Stress-rest” | 0.98 | 0.19 | 2.65 (1.82-3.87) | <0.001 |
| Any ischemia by “Stress-LGE (>0%)” | 0.91 | 0.019 | 2.48 (1.71-3.61) | <0.001 |
| Any ischemia by “Stress-LGE (>75%)” | 0.97 | 0.19 | 2.65 (1.82-3.86) | <0.001 |
| Ischemia burden by “Stress-rest” | 0.11 | 0.03 | 1.11 (1.05-1.12) | <0.001 |
| Ischemia burden by “Stress-LGE (>0%)” | 0.11 | 0.03 | 1.12 (1.5-1.19) | 0.0004 |
| Ischemia burden by “Stress-LGE (>75%)” | 0.10 | 0.03 | 1.10 (1.05-1.16) | <0.001 |
| Any infarction by LGE | 0.99 | 0.20 | 2.69 (1.82-3.98) | <0.001 |
| Infarction burden | 0.15 | 0.05 | 1.16 (1.06-1.27) | 0.002 |
| LVEF | -0.19 | 0.01 | 0.98 (0.96-1.01) | 0.98 |

**Table 4.**
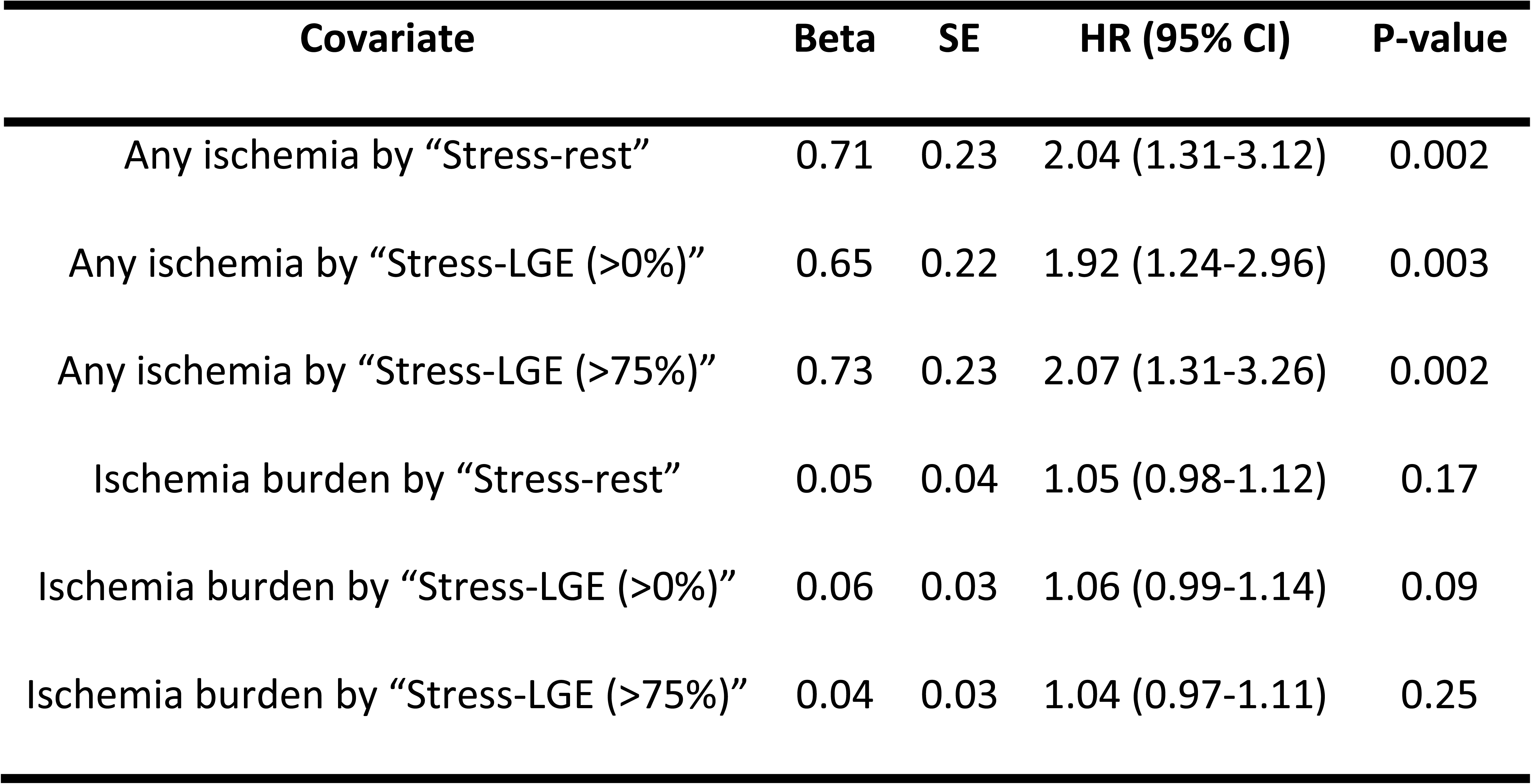
Multivariable Cox regression of “Stress-rest” and “Stress-LGE” strategies corrected for LVEF and any infarction by LGE. Beta is the coefficient of the model, SE the Standard Error, and HR the Hazard Ratio.

## DISCUSSION

In this study we have performed a new exploratory analysis of data from 666 patients from CE-MARC with complete stress perfusion, rest perfusion, LGE and QCA data. The optimum definition of inducible ischemia was “Stress-LGE (>75%)”; the presence of a stress-induced perfusion defect without transmural infarction. This definition had higher diagnostic accuracy than both “Stress-LGE (>0%)” and “Stress-rest”, the latter being validated in the original CE-MARC analysis.

In patients with MI, use of “Stress-rest” analysis as the threshold for classifying a segment as ischemic increased the ischemia burden compared with the “Stress-LGE (>0%)” definition, likely reflecting the incorrect classification of sub-segmental peri-infarct ischemia as infarction in the latter. Finally, a positive test was associated with adverse outcomes on long term follow up regardless of whether “Stress-rest” or “Stress-LGE” analysis was used. These results support the use of a “Stress-LGE” image analysis without the need for acquisition of rest perfusion imaging.

### Diagnostic accuracy of imaging protocols

In this study we have shown that removing rest perfusion from the imaging analysis does not adversely impact diagnostic accuracy measured against QCA, and in fact the diagnostic accuracy can be improved when only transmurally infarcted segments are classified as non-ischemic. The sensitivity and specificity were not significantly different between the two analysis methods (“Stress-rest” 73.6% and 93.1% and “Stress-LGE (>75%)” 73.6% and 93.1% respectively). It should be noted that the sensitivity in this analysis was lower than in the main trial where a positive test was defined by multiparametric findings including LGE and wall motion score^3^.

The data from CE-MARC is relatively unique in that all patients had both QCA and stress perfusion CMR. It is therefore ideal to validate the diagnostic accuracy of “Stress-only” imaging. Previous studies have examined the utility of “Stress-only” protocols. Rijlaarsdam-Hermsen et al. reported CMR findings from 642 consecutive patients with chest pain and a non-zero coronary artery calcium score^4^. They reported that “Stress-only” adenosine CMR was associated with a 91% sensitivity and a 99% specificity for the identification of coronary artery disease and showed incremental diagnostic benefit to coronary artery calcium scoring alone. However, invasive angiography was not mandated and was only performed if the stress-CMR was positive, introducing a major selection bias to the data. Additionally, a previous sub-study of CE-MARC, comparing quantitative myocardial perfusion assessments to visual analysis, found no benefit to inclusion of the rest perfusion data to the analysis for the detection of significant CAD^13^.

### Refining diagnostic accuracy stress-only imaging

Only 18.6% of patients in this study had MI by LGE imaging but the classification of infarcted segments made a large difference to both diagnostic accuracy and ischemia burden. We have shown that for the highest diagnostic accuracy in segments with both infarct and stress perfusion defects, only segments with transmural infarction should be classified as infarct, whereas all segments with sub-endocardial infarction and stress perfusion defects should be classified as ischemic. The fact that this definition of “Stress-LGE” imaging outperforms “Stress-rest” imaging likely reflects the higher spatial resolution of LGE imaging compared to rest perfusion imaging, and therefore the more accurate delineation of peri-infarct ischemia.

In the analysis we have only reported our findings segmentally. With advances in perfusion and LGE imaging, there are opportunities to improve the in-plane spatial resolution and increase the number of slices acquired in stress imaging. These advances will allow differentiation of infarct and peri-infarct ischemia on a subsegmental level from a “Stress-only” scan leading to further improvements in diagnostic accuracy.

Villa et al. examined the impact of reporter experience on diagnostic accuracy. The main determinant of diagnostic accuracy was level of training. Rest-perfusion imaging did not improve diagnostic accuracy, although it did contribute to higher confidence in the results, particularly with the addition of quantitative perfusion maps which are particularly helpful in the exclusion of balanced ischemia^14^. Inclusion of rest-perfusion imaging could also still have a role when stress-perfusion imaging is affected by artefact, although this is equally likely to persist in rest-imaging.

### Prognostic importance of a positive test

In this analysis of CE-MARC the presence of inducible ischemia by either "Stress-rest” or “Stress-LGE (>75%)” was associated with MACE over median 6.8 years follow up (hazard ratios for both 2.65). There is limited data from other studies on the prognostic potential of “Stress-only” imaging but there are several studies which have shown the prognostic importance of inducible ischemia detected on “Stress-rest” CMR^15–17^.

SPINS (Stress CMR Perfusion Imaging in the United States) is a large multicenter retrospective registry of patients undergoing stress perfusion CMR for the evaluation of chest pain. SPINS included data from 2349 patients followed up for median of 5.4 years^18^. The protocol did not mandate rest-perfusion imaging (although this was standard practice during the period of recruitment from 2008-2013). In SPINS the presence of inducible ischemia was associated with increased risk of MACE (3.30; 95% CI: 2.67–4.08, <0.0001) which is comparable to the findings of our study.

The landscape of coronary intervention for stable angina has changed significantly over the last two decades, with the role for PCI as the initial strategy diminishing. The ISCHEMIA trial^21^ showed no additional benefit to PCI in the context of demonstrable ischemia and that a more conservative medical strategy could be safely employed. CE-MARC 2^10^ showed that stress CMR was less likely to result in invasive angiography than the contemporaneous NICE guidelines with no difference in MACE rates. MR-INFORM^23^ showed that stress CMR guided care resulted in lower rates of PCI than fractional flow reserve (FFR) guided care, with similar rates of MACE. Stress CMR therefore has strong evidence for a role as a gatekeeper test for invasive angiography, and we advocate based on the data presented here this can safely be done without rest perfusion.

### Clinical benefits of a Stress-LGE imaging protocol

There are several benefits of a “Stress-LGE” imaging strategy. CMR scan time could potentially be significantly reduced if rest perfusion imaging is not acquired, particularly if other rapid image acquisition techniques are used^25,26^, therefore widening access, decreasing cost, and reducing waiting times. Alternatively, the saved time could be used to acquire flow or mapping sequences, that have greater prognostic relevance than rest-perfusion. Given that stress CMR is recommended in both US chest pain^1^ and ESC chronic coronary syndrome^27^ guidelines, increased demand and streamlined protocols will likely increase capacity and availability. There is also the option to repeat stress imaging without concerns about total contrast dose if the first stress imaging is non-diagnostic. Finally, a “Stress-LGE” strategy is ideal for Regadenoson which, preferably because of its long half-life, requires left ventricular volumes and rest-imaging to be acquired before its administration^28^. For the purposes of streamlining, LV volumes can be acquired after regadenoson stress, making the scan even shorter (but acknowledging that LVEF may be slightly increased)^28^.

### Study Limitations

In addition to the previously reported limitations of CE-MARC^3,13^, in this analysis we have performed the separate “Stress-rest” and “Stress-LGE” analyses from a single scan. We have used the original segmental analysis of CE-MARC for this study where the scan was reported in its entirety. It is possible therefore that unconscious bias may have affected the segmental reporting, although this may be offset by the consensus methodology used. Perfusion and LGE imaging were not matched for slice location although all 16 segments were covered by both imaging techniques. QCA was used as the reference standard in CE-MARC rather than FFR, as this was standard practice at the time of data acquisition during CE-MARC which predated the major FFR trials. The proportion of patients in this analysis with evidence of prior MI on LGE imaging was relatively small at 19%, which might limit statistical power.

## Conclusions

The optimum definition of inducible ischemia was the presence of a stress-induced perfusion defect without transmural infarction. This definition improved the diagnostic accuracy compared to the “Stress-rest” analysis validated in CE-MARC without the need for rest-perfusion imaging. AHA/ACC/ASE/CHEST/SAEM/SCCT/SCMR chest pain guidelines recommend stress perfusion CMR in patients with stable chest pain^1^, and these results are reassuring that the absence of ischemia by either analysis strategy confers favorable long-term prognosis.

## Data Availability

The datasets generated and analysed during the current study are not publicly available. Data are available from the corresponding author upon reasonable request.

## ABBREVIATIONS AND ACRONYMS

CAD: Coronary Artery Disease
CE-MARC: Clinical Evaluation of MAgnetic Resonance imaging in Coronary heart disease
CMR: Cardiovascular Magnetic Resonance
FFR: Fractional Flow Reserve
LGE: Late Gadolinium Enhancement
LVSD: Left Ventricular Systolic Dysfunction
MACE: Major Adverse Cardiac Events
MI: Myocardial Infarction
MPS-SPECT: Myocardial perfusion scintigraphy using single-photon emission computerized tomography
QCA: Quantitative coronary angiography
ROC: Receiver Operator Curve
SCMR: Society for Cardiovascular Magnetic Resonance

---

**Central Illustration.**
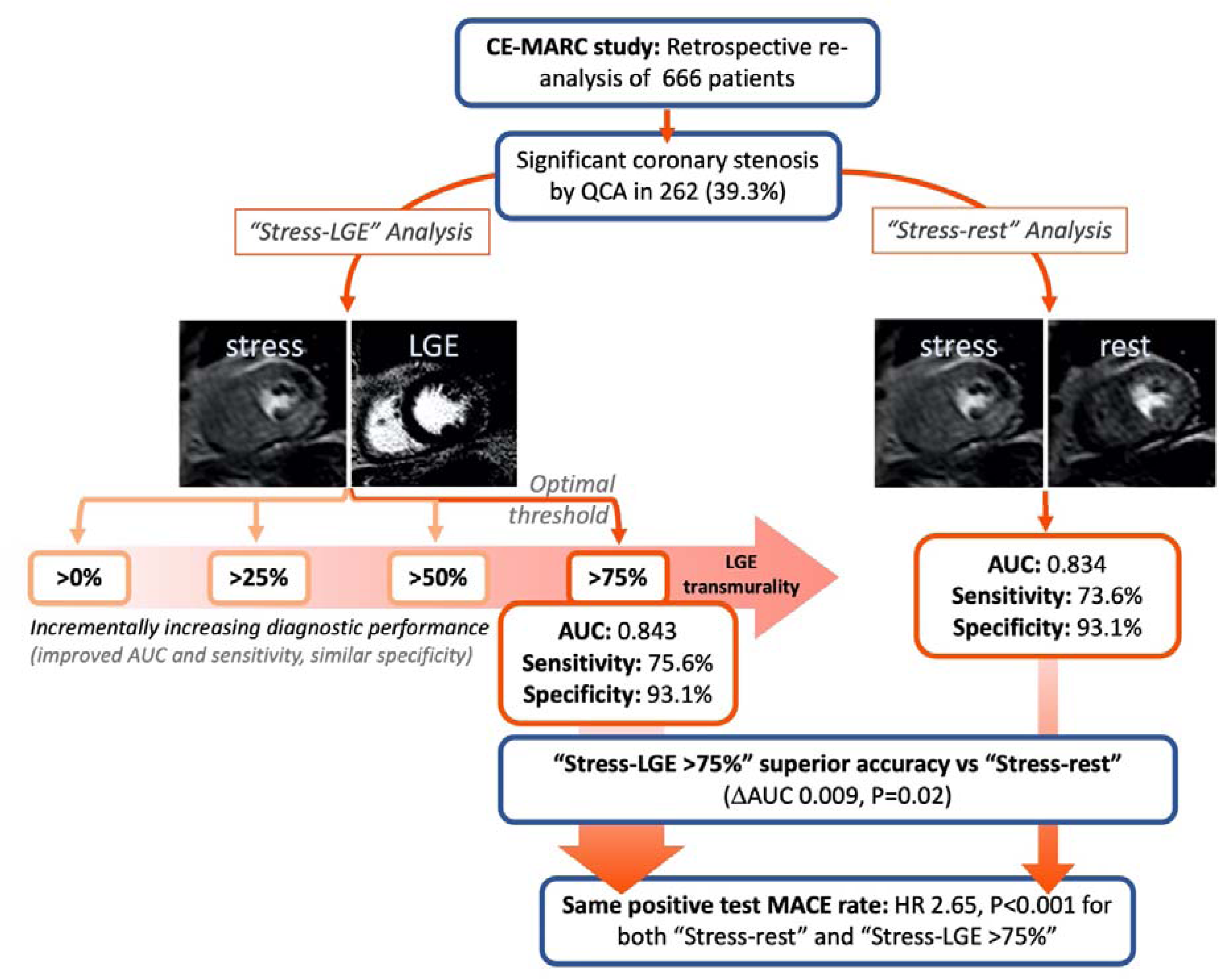
Summary of “Stress-LGE” reanalysis of the CE-MARC study and its main findings.

